# Comparison of Masimo Rad-67 SpHb non-invasive hemoglobin monitoring device with complete blood count measurement for use in pregnancy: An observational multi-site cohort study

**DOI:** 10.64898/2025.12.14.25342241

**Authors:** Fouzia Farooq, Yipeng Wei, Qing Pan, Abigale Proctor, Sasha G. Baumann, Kevin Kasadhe, Harun Owour, Caleb Sagam, Azqa Mazhar, Nida Yazdani, Humphrey Mwape, Augustine Tunga, Victor Akelo, Christopher Mores, Margaret P. Kasaro, Muhammad Imran Nisar, Wilbroad Mutale, M. Bridget Spelke, Emily R. Smith, Zahra Hoodbhoy, the Pregnancy Risk, Infant Surveillance, and Measurement Alliance (PRISMA) Consortium

## Abstract

**Background:** Anemia remains a major health concern, particularly during pregnancy. However, blood draws, laboratory capacity, processing time, and equipment cost required for hemoglobin testing to diagnose anemia can be prohibitive. The Total Hemoglobin SpHb Rad-67 Pulse CO-Oximeter offers a non-invasive, portable, and affordable point-of-care alternative. We aimed to validate SpHb against complete blood count (CBC) in a pregnant and postpartum population.

**Methods:** This was a substudy of the Pregnancy Risk, Infant Surveillance, and Measurement Alliance (PRISMA) Maternal and Newborn Health Study. A total of 2,700 participants in Zambia, Kenya, and Pakistan provided hemoglobin measurements both by CBC and the Rad-67 Masimo SpHb device at four visits during pregnancy and at six-weeks postpartum. We assessed agreement between SpHb and CBC and used mixed models to identify factors that explained or influenced differences between the two methods.

**Results:** We found the mean hemoglobin measurement by SpHb (12.8±1.6 g/dL) was higher than by CBC (11.0±1.6 g/dL). However, this overall positive bias masked systematic misclassification at the extremes: SpHb overestimated values among women with very low hemoglobin (Mean Difference (MD) =-3.95; 95%CI-4.28,-3.62) and underestimated values at very high hemoglobin levels (MD = 2.44; 95%CI 2.14, 2.74), even after adjustment. Using CBC, 48% of observations were classified as anemic (<11 g/dL), compared to 9% by SpHb; conversely, just 7% of CBC readings fell within 13-15 g/dL, compared to 38% by SpHb. Agreement metrics consistently showed poor concordance between the two methods.

**Conclusions:** SpHb systematically overestimated hemoglobin on average and showed poor agreement with CBC, particularly at clinically relevant extremes. Until greater accuracy of SpHb is demonstrated in this population, hemoglobin testing with laboratory-based methods is recommended to inform clinical decision-making in pregnancy.

**AUTHOR SUMMARY:** Anemia is a major global health challenge linked with maternal morbidity, adverse birth outcomes, and impaired infant development. Accurate and accessible hemoglobin testing is critical for timely anemia diagnosis and treatment. The gold standard for hemoglobin testing is complete blood count using venous blood; however, this method requires laboratory infrastructure, blood sample collection, and trained personnel, limiting feasibility in low-resource and rural settings where anemia burden is highest. We evaluated the accuracy of the Masimo Total Hemoglobin SpHb® measured by Rad-67® Pulse CO-Oximeter®: a device that measures hemoglobin via an optical sensor placed on the patient’s finger. We found the mean hemoglobin measurement by SpHb (12.8±1.6 g/dL) was higher than by CBC (11.0±1.6 g/dL).We found that the Masimo device consistently overestimated hemoglobin at very low levels and underestimated at very high levels. Masimo SpHb classified 9% of women as anemic (i.e. hemoglobin <11 g/dL), compared to 48% using the gold standard method. The Masimo SpHb was even less accurate for women later in gestation, living with HIV, and who reported using betelnut, tobacco, or smoking. Device improvements or software-based correction factors are needed to improve performance of the Total Hemoglobin SpHb Rad-67 Pulse CO-Oximeter before it can be used to measure hemoglobin in pregnancy.

## INTRODUCTION

Globally, 36% of pregnant women and 31% of women of reproductive age (15 to 49 years) suffer from anemia [1]. In Kenya, Pakistan, and Zambia, the rates of anemia in women aged 15 to 49 are higher than the global average, at 40%, 39%, and 37%, respectively [2]. Identifying and treating anemia is critical for maternal and newborn health, as moderate and severe anemia in pregnancy is associated with increased risk of adverse outcomes including preterm birth, severe postpartum hemorrhage, maternal death, stillbirth, and fetal growth restriction [3]. Early detection and effective management can therefore play a pivotal role in improving pregnancy outcomes and reducing preventable morbidity and mortality. Complete blood count (CBC) is the gold standard laboratory test for hemoglobin (Hb) measurement; this test relies on an invasive venous blood sample collected from the patient and is run on a hematology analyzer by a trained technician [4]. A non-invasive method for measuring hemoglobin could significantly enhance patient care by providing faster results, minimizing exposure to potential biohazards, and reducing discomfort for patients [5]. In resource-limited settings, where venous blood draws and laboratory testing may not be routinely available, there is a need for low-cost, accurate, point-of-care hemoglobin measurement strategies to diagnose anemia in pregnancy.

Point-of-care devices like the HemoCue can increase testing access by removing barriers such as cost of equipment, cold chain, and reliance on skilled personnel [6]. Point-of-care devices also give rapid readings to allow providers to utilize results in clinical decision-making within the same visit. However, these devices are still considered invasive due to the required finger prick for blood sample collection. The Masimo saturation of peripheral hemoglobin (SpHb) devices offer a fully non-invasive point-of-care alternative, using pulse oximetry to measure hemoglobin levels via an optical sensor on the patient’s finger. While the device has received US Food and Drug Administration approval for use in the general population, it has not been approved for use in pregnancy [7]. This gap reflects a broader lack of clinical evidence and scientific consensus regarding the device’s accuracy for diagnosing anemia in pregnant populations. Nonetheless, its non-invasive nature and the lack of blood sampling or laboratory testing make the device particularly promising for use in low-resource and remote settings.

Generally, the reported accuracy of point-of-care devices for Hb measurement has been inconsistent. Some studies have supported comparable Hb measurement between point-of-care devices and the CBC gold standard, with accurate detection of Hb in adult populations and a similar degree of bias and standard deviation [8–10]. Others report limited evidence on device accuracy or do not specifically evaluate Masimo SpHb. Our 2024 meta-analysis of non-invasive hemoglobin testing using the Radical-7 pulse co-oximetry in surgical patients found no clinically significant difference between CBC and non-invasive measurements overall, though CBC values were slightly higher in adults and lower in children [11]. A 2015 systematic review of Masimo SpHb and HemoCue performance found that SpHb had lower precision and wider 95% limits of agreement than the HemoCue; similarly, a study in pregnant patients found substantial variability in SpHb bias and limits of agreement, indicating inconsistent accuracy [12, 13]. In a study in perioperative patients, the Masimo Rad-67 showed some accuracy, but with greater differences at lower values [14]. Another Masimo device, the Pronto 7, has been used in the adult population as well as healthy children with a wide sensitivity range of 54-93% [15, 16] and a specificity of 75-95% for detection of anemia [17, 18]. Research on these devices in pregnancy has been limited. One study on pregnant women in Kenya showed high specificity and positive predictive value with the Masimo SpHb, but inconsistency in Hb values, while another in postpartum women showed a consistent overestimation in Hb values [19, 20]. Further research is needed to determine the accuracy of Masimo SpHb for hemoglobin testing to diagnose anemia in pregnancy.

The assessment of Masimo Total Hemoglobin SpHb® for Hb measurement accuracy compared to gold standard was conducted as part of the Pregnancy Risk, Infant Surveillance, and Measurement Alliance (PRISMA) Maternal and Newborn Health Study. PRISMA is a population-based, open-cohort study evaluating the associations between pregnancy risk factors and adverse pregnancy outcomes across six study sites in five countries in sub-Saharan Africa and South Asia. The goals of PRISMA are described in the study protocol, available on Open Science Foundation [21]. Anemia is a secondary outcome of the PRISMA Maternal and Newborn Health Study and Hb is measured at every scheduled study visit throughout pregnancy and through one year postpartum. For the purpose of the present validation study, sites in Kenya, Pakistan, and Zambia collected Hb data using CBC and additionally with the Masimo Rad-67 SpHb device at select timepoints.

The overarching aim of this study was to evaluate the accuracy of total Hb measurements using the Masimo SpHb compared to gold standard CBC Hb measurement. Specific study aims were as follows: (1) Estimate the amount of deviation between longitudinal SpHb and CBC values; (2) Assess agreement level between SpHb and CBC values among binary (healthy versus anemic) and ordinal (mild, moderate, severe anemia, versus normal) measures; and (3) Describe sociodemographic and clinical factors affecting SpHb and CBC deviation.

## MATERIALS AND METHODS

### Study Design

Data used in the present analysis was collected between September 2022 and January 2024 at three study sites in Karachi, Pakistan; Kisumu and Siaya, Kenya; and Lusaka, Zambia. A detailed summary of study design and recruitment methods is presented in Farooq et al., 2024 [24]. This analysis restricted inclusion to the first 900 consecutively enrolled women per site (total N=2,700) with complete identifiers and no inconsistencies across linked data.

### Sample Size

The sample size of 900 participants per site gave us a 90% power to conduct site specific analyses. These repeated-measures provided greater than 90% power for the primary analysis.

This is supported by preliminary calculations which showed that, for a simple cross-sectional design, only 19 pairs of SpHb and CBC measurements were needed to achieve 90% power. Details of the sample size calculations and statistical rationale are previously published in Farooq et al., 2024 [24].

### Participants

Participants were women enrolled in the ongoing PRISMA Maternal and Newborn Health Study. Eligibility criteria for PRISMA study enrollment were as follows: lives within the study catchment area with no plans to permanently relocate during pregnancy or within one year postpartum; meets minimum age requirement for consenting in country of residence; viable intrauterine pregnancy less than 20 weeks gestation determined by ultrasound; and provides informed consent. Participant-level data were drawn from case report forms and linked using unique individual and pregnancy IDs. Variables were standardized and cleaned across datasets. Records from duplicate IDs were excluded and, for individuals with multiple pregnancies, only data from the first pregnancy were retained. All participant data were de-identified prior to analysis to ensure privacy and confidentiality. Selected women provided hemoglobin estimates by both CBC and Masimo SpHb at five scheduled PRISMA study visits: at less than 20 weeks gestation, 20 weeks gestation, 28 weeks gestation, 36 weeks gestation, and 6 weeks postpartum (Figure 1).

**Figure 1.**
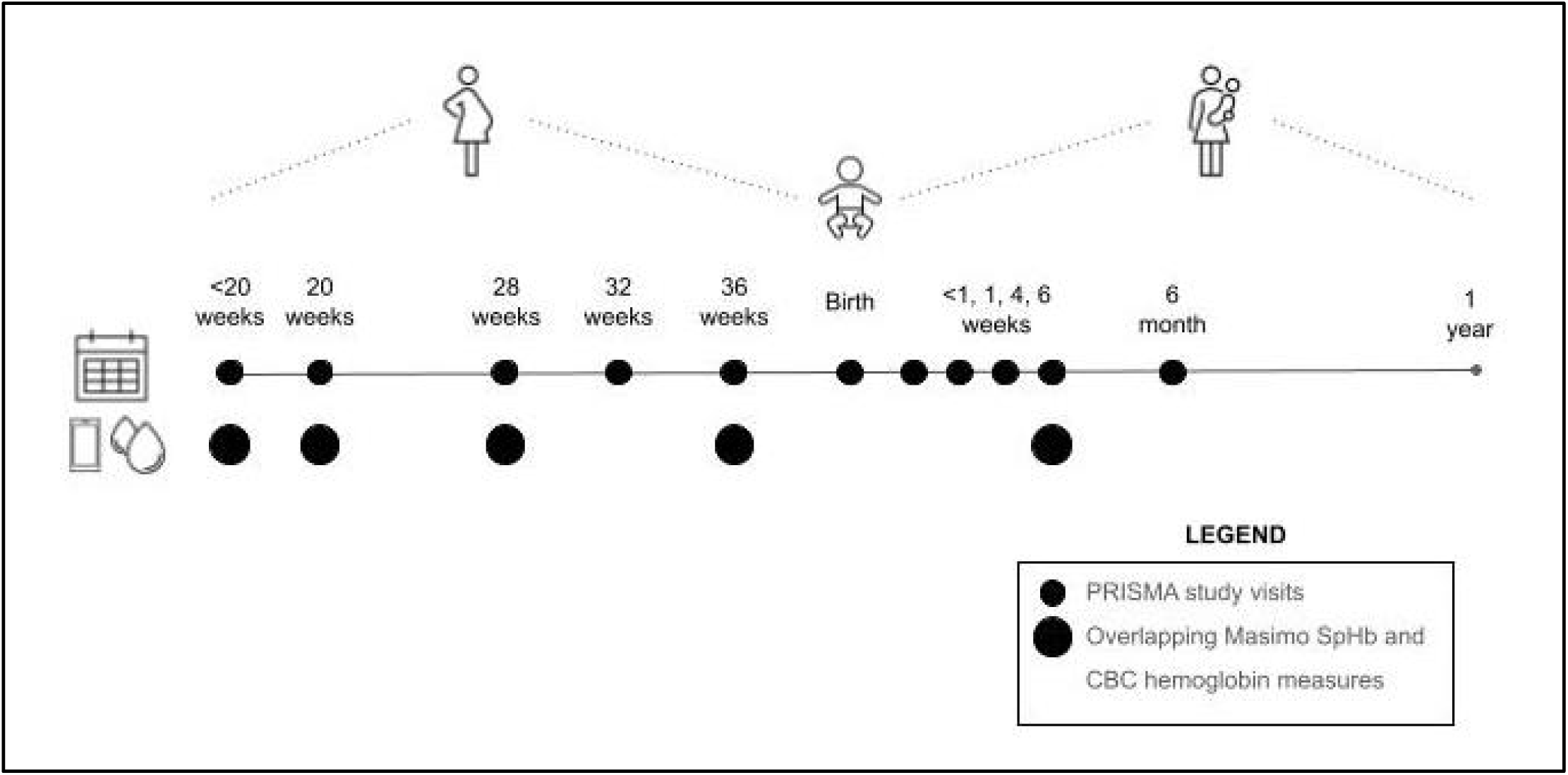
Nested prospective cohort study design

### Test Methods

Maternal SpHb hemoglobin was assessed using Masimo non-invasive Rad-67 at five study visits (<20 weeks, 20 weeks, 28 weeks, 36 weeks gestation, and 6 weeks postpartum). Corresponding venous blood samples were processed at site laboratories for CBC using the following hematology analyzers: Beckman Coulter Ac•T 5diff CP (Cap Pierce) in Kenya, the Sysmex XN-1000 in Pakistan, and the Beckman Coulter DxH520 in Zambia. Results were monitored via two external quality assessment platforms: UK-NEQAS (United Kingdom National External Quality Assessment Scheme) and CAP (College of American Pathologists). Measurement dates were aligned to ensure that SpHb and CBC were obtained at or within 7 days of the scheduled visit date by gestational age. CBC values were adjusted for altitude using the World Health Organization 2024 *Guideline on haemoglobin cutoffs to define anemia in individuals and populations*; as such, a subtraction of 0.8 g/dL was applied to all values for participants in Kenya and Zambia [25].

### Analysis

The primary outcome was the difference in SpHb and CBC Hb values at each time point and overall. Specific analytical objectives as follows:

1. Assess agreement between SpHb and CBC Hb on a continuous scale.
2. Assess the degree of agreement between SpHb and CBC Hb on a categorical scale.
3. Assess factors affecting differences between SpHb and CBC Hb.

To evaluate the agreement and correlation between SpHb and CBC Hb measurements at each visit, intraclass correlation coefficient (ICC) and Pearson correlation coefficient (PCC) were used, followed by the Bland-Altman analysis. ICCs were calculated with the *irr* R package for each study site separately to assess the absolute agreement between SpHb and CBC hemoglobin levels using a two-way mixed-effects model with absolute agreement and single measurement assumption. The PCC were also computed separately for each site using pairwise complete observations using the *cor()* function in R. The ICC estimates represent the proportion of total variance, both within subjects and between the SpHb and CBC methods, in hemoglobin levels attributable to differences between measurement methods. Pearson’s r provides a measure of the strength and direction of the linear relationship between SpHb and CBC values. Both methods measure the similarity between hemoglobin levels from SpHb and CBC hemoglobin measured on the same patient at each time point. The Bland-Altman plots assess the agreement between the SpHb and CBC measurements by plotting the difference between a pair of SpHb and CBC values over the average of the two values, with significant differences from zero indicating systematic over-or underestimates of the hemoglobin values.

To assess the degree of agreement between SpHb and CBC Hb on a categorical scale, anemia status was evaluated as a binary measure using Positive Percent Agreement (PPA) and Negative Percent Agreement (NPA), and Cohen’s kappa. PPA and NPA were employed to measure the agreement between the SpHb and CBC measures in order to identify binary anemia cases. Four different hemoglobin cutoffs were applied to define anemia and elevated hemoglobin status, including Hb <11 g/dl or Hb >13 g/dl; Hb <11 g/dl or Hb >15 g/dl; Hb <11 g/dl only; Hb >15 g/dl only. To estimate the PPA and NPA, we used generalized linear mixed-effects models with a logit link function and the delta method to compute the variances of PPA and NPA [26]. Additional analyses, including Cohen’s kappa and McNemar’s tests, were conducted to evaluate the reliability and agreement between the two methods on a binary scale, as described previously [24].

To assess the degree of agreement between SpHb and CBC hemoglobin on an ordinal scale, weighted Cohen’s kappa and Harrel’s concordance index (C-Index) were used. Weighted Cohen’s kappa quantifies interrater agreement on ordinal anemia severity categories (normal, mild, moderate, and severe anemia, and high and very high hemoglobin). Agreement was weighted based on the extent of discrepancy between SpHb and CBC classifications using different weights. Harrel’s concordance index (C-Index) qualifies the proportion of concordant pairs among all comparable pairs in the dataset. A pair is considered concordant if both SpHb and CBC measurements rank anemia status in the same order.

We defined anemia status based on the 2024 World Health Organization hemoglobin thresholds for anemia in pregnant and non-pregnant adults [25] During pregnancy, thresholds are trimester-specific: First and third trimesters: no anemia (≥11.0 g/dL), mild anemia (10.0 - 10.9 g/dL), moderate anemia (7.0 - 9.9 g/dL), severe anemia (<7.0 g/dL); Second trimester: no anemia (≥10.5 g/dL), mild anemia (9.5 - 10.4 g/dL), moderate anemia (7.0 - 9.4 g/dL), severe anemia (<7.0 g/dL).

Postpartum (i.e. non-pregnant) anemia categories are as follows: No anemia (≥12.0 g/dL), mild anemia (11.0 - 11.9 g/dL), moderate anemia (8.0 - 10.9 g/dL), severe anemia (<8.0 g/dL).

The differences in hemoglobin levels between SpHb and CBC were evaluated in the context of a broad set of maternal risk factors known to influence anemia during pregnancy [22, 23]. Univariate and multivariate linear mixed models were fitted to test candidate risk factors versus the difference between pairs of SpHb and CBC measurement values. Model diagnosis, such as variance inflation factor and penalized regression for variable selection, were conducted. The following clinical and sociodemographic risk factors were assessed to determine the difference in SpHb and CBC measurements: maternal age of <18 years; tobacco, betelnut use and/or cigarette smoking; body mass index; gestational age at enrollment; gestational age by trimester; HIV status at enrollment; malaria status at any visit during pregnancy; study site; maternal anemia status; plasma volume expansion; mid-upper arm circumference; and ferritin. All analyses were conducted in R version 3.2.1.

## RESULTS

### Participants

A sample of 2,700 women (900 from each study site) were included in this analysis. Table 1 summarizes the baseline characteristics of the included women. Overall, the average maternal age was 27.1±5.7 years with a mean gestational age of 13.4±3.7 weeks at enrollment. Seventy nine percent of women had some education and 88% of women were married. Tobacco use, betelnut use or smoking was reported at 13% (highest in Pakistan at 39% driven by betelnut use). Approximately half (51%) of participants had a normal early pregnancy body mass index of 18 to <25 kg/m^2^ at the time of enrollment (<20 weeks gestation) with notable between-site differences in underweight and overweight trends.

**Table 1.** Baseline characteristics of included women overall and stratified by study site.

| Characteristic | Overall<br>N = 2,700<sup>1</sup> | Kenya<br>N = 900<sup>1</sup> | Pakistan<br>N = 900<sup>1</sup> | Zambia<br>N = 900<sup>1</sup> |
| --- | --- | --- | --- | --- |
| Gestational age at enroll (weeks) | 13.4 (3.7) | 13.8 (3.7) | 12.1 (3.6) | 14.2 (3.6) |
| Maternal age (years) | 27.1 (5.7) | 26.9 (5.2) | 26.5 (5.9) | 27.9 (6.0) |
| Maternal age (<18 years) | 31 (1.1%) | 0 (0%) | 11 (1.2%) | 20 (2.2%) |
| Ever attended school | 2,136 (79%) | 895 (100%) | 357 (40%) | 884 (99%) |
| Wealth quintile |  |  |  |  |
| Lowest quintile | 499 (19%) | 167 (19%) | 178 (20%) | 154 (17%) |
| Second quintile | 478 (18%) | 151 (17%) | 151 (17%) | 176 (20%) |
| Third quintile | 510 (19%) | 179 (20%) | 161 (18%) | 170 (19%) |
| Fourth quintile | 635 (24%) | 191 (21%) | 218 (24%) | 226 (25%) |
| Top quintile | 575 (21%) | 211 (23%) | 190 (21%) | 174 (19%) |
| Marital status |  |  |  |  |
| Married | 2,376 (88%) | 747 (83%) | 898 (100%) | 731 (81%) |
| Not married but cohabitating | 13 (0.5%) | 13 (1.4%) | 0 (0%) | 0 (0%) |
| Divorced/separated | 25 (0.9%) | 9 (1.0%) | 0 (0%) | 16 (1.8%) |
| Widowed | 5 (0.2%) | 4 (0.4%) | 0 (0%) | 1 (0.1%) |
| Single/never married | 278 (10%) | 126 (14%) | 0 (0%) | 152 (17%) |
| Parity (livebirths & stillbirths) |  |  |  |  |
| 0 | 739 (27%) | 256 (29%) | 264 (29%) | 219 (24%) |
| 1 | 723 (27%) | 273 (30%) | 212 (24%) | 238 (26%) |
| 2+ | 1,234 (46%) | 368 (41%) | 423 (47%) | 443 (49%) |
| Tobacco, betelnut or smoking | 352 (13%) | 3 (0.3%) | 346 (39%) | 3 (0.3%) |
| BMI |  |  |  |  |
| <18.5 kg/m<sup>2</sup> | 294 (11%) | 47 (5.2%) | 216 (24%) | 31 (3.4%) |
| 18-<25 kg/m<sup>2</sup> | 1,375 (51%) | 519 (58%) | 460 (51%) | 396 (44%) |
| 25-<30 kg/m<sup>2</sup> | 647 (24%) | 218 (24%) | 157 (17%) | 272 (30%) |
| 30+ kg/m<sup>2</sup> | 381 (14%) | 114 (13%) | 66 (7.3%) | 201 (22%) |
| MUAC <20 weeks (≤23 cm) | 27.1 (4.5) | 28.1 (4.2) | 25.0 (4.3) | 28.2 (4.3) |
| HIV at enrollment | 285 (11%) | 116 (13%) | 1 (0.1%) | 168 (19%) |
| Malaria at enrollment | 134 (5.0%) | 127 (14%) | 1 (0.1%) | 6 (0.7%) |

<sup>1</sup>Mean (SD); n (%)
BMI: Body Mass Index
MUAC: Mid upper arm circumference

### Test Results

There was a high degree of protocol compliance, with overall follow-up visit completion of 89.6%. Data completeness (Table S1), however, varied by visit and declined over subsequent study visits, with the highest completion at the enrollment visit and lowest at six weeks postpartum.

Masimo SpHb consistently produced higher hemoglobin readings than CBC measurements across study sites at all timepoints. Pakistan had an overall mean SpHb measurement of 12.2±1.3 g/dL, compared to a mean CBC measurement of 10.9±1.5 g/dL. In Kenya mean SpHb and CBC hemoglobin were 13.0±1.2 g/dL and 10.5±1.6 g/dL, respectively. Zambia had a mean SpHb and mean CBC hemoglobin of 13.1±1.8 g/dL and 11.4±1.5 g/dL, respectively. The overall mean SpHb measurement across all sites was 12.8±1.6 g/dL and the overall mean CBC hemoglobin measurement was 11.0±1.6 g/dL, demonstrating a consistent positive bias in SpHb readings.

Visual inspection using the Bland-Altman plots showed that while most hemoglobin measurements from the two methods (SpHb and CBC) on the same sample fell within the 95% limits of agreement (Figure 2), the wide spread of these limits indicates substantial variability between the two methods. Although the mean difference across sites and time points was close to zero, it varied in both direction and magnitude depending on the country and gestational age at time of measurement.

**Figure 2.**
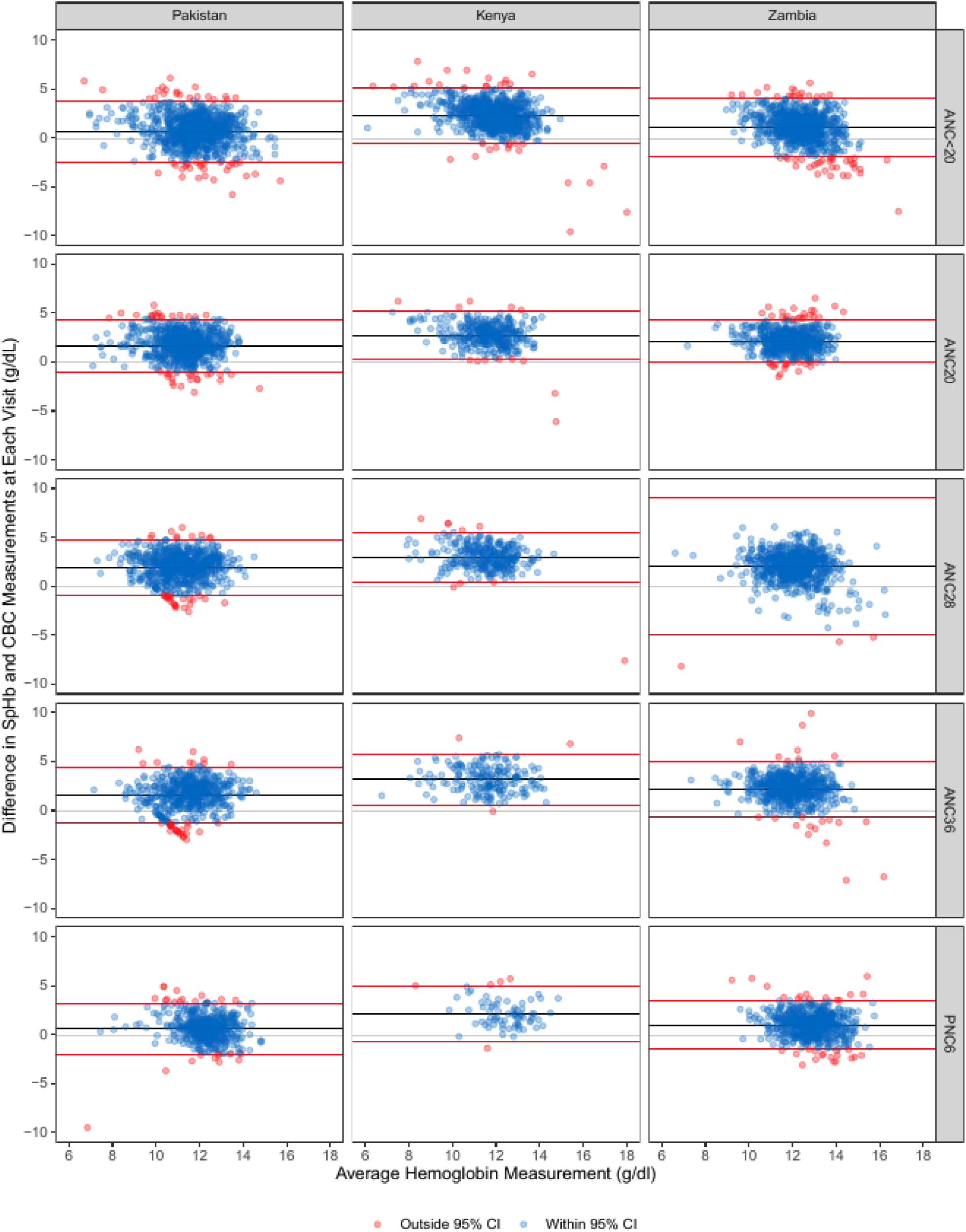
Bland-Altman plots showing the difference between SpHb and CBC values against their average, stratified by study sites and across five antenatal (ANC) and postnatal (PNC) care time points (defined by weeks).

Notably, the greatest variability between methods was observed among the Zambia sample at the 28 week antenatal visit. A slight positive bias (SpHb>CBC) was observed around an average hemoglobin value of 12 g/dL, and a slight negative bias at ≥12 g/dL, particularly at enrollment.

These patterns suggest measurement inconsistency rather than reliable agreement. This is further supported by the low ICCs and PCCs observed across all sites and timepoints (Table 2).

**Table 2.**
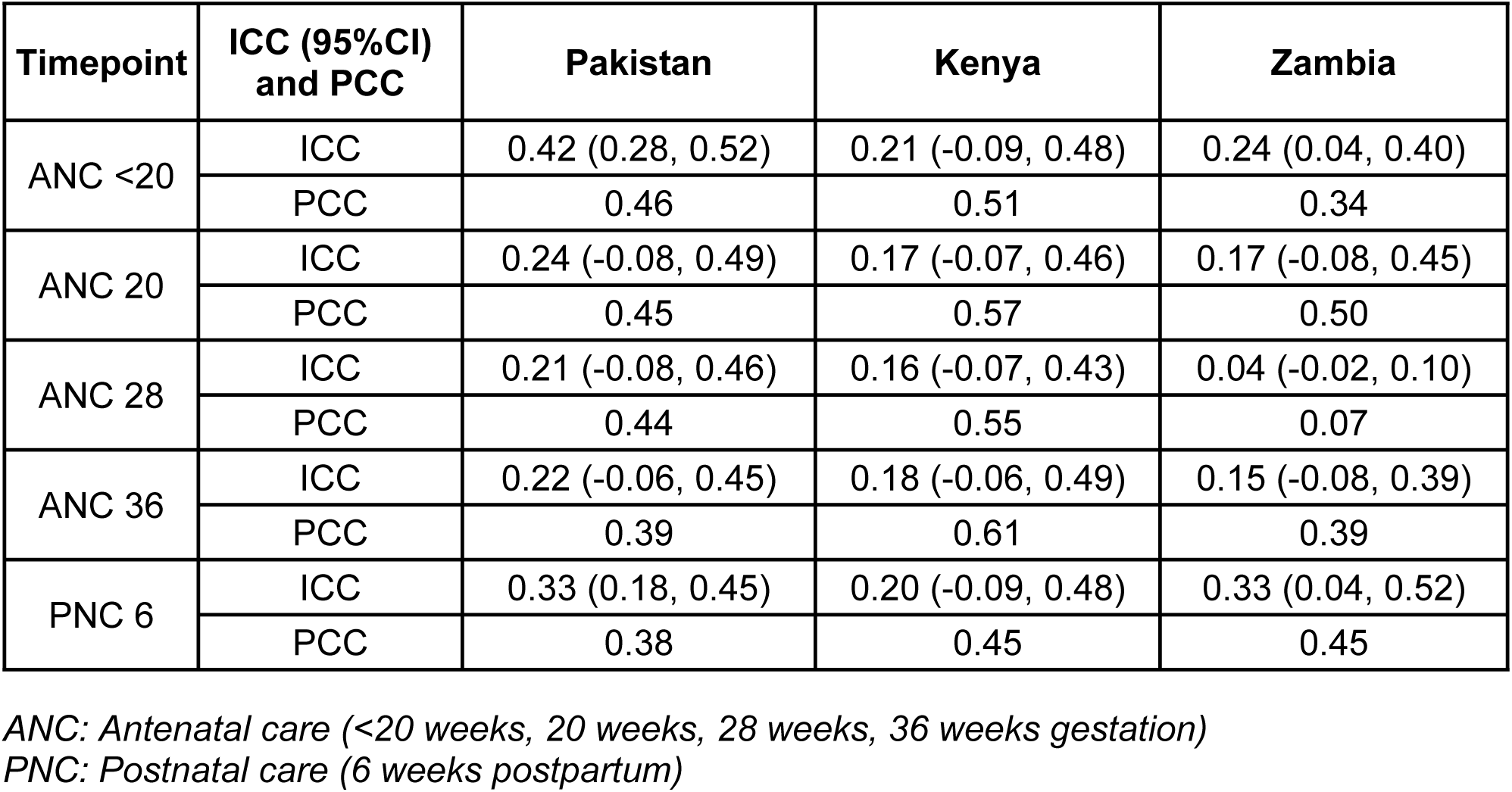
Intraclass correlation coefficient (ICC) and Pearson correlation coefficient (PCC) between CBC and SpHB at each time point for each site.

ICCs ranged from 0.04 (95%CI-0.02, 0.10) at the 28 week visit in Zambia to 0.42 (95%CI 0.28, 0.52) at the enrollment visit in Pakistan. PCCs were also comparable across all three sites although slightly higher, ranging from 0.07 at the 28 week visit in Zambia to 0.61 at the 36 week visit in Kenya, indicating poor overall agreement between the two measurement methods.

Considering the distribution within different clinical thresholds of hemoglobin levels, Masimo SpHb tended to overestimate low hemoglobin values indicative of anemia and underestimate normal or elevated hemoglobin levels. Specifically, 48.3% of the observations by CBC found hemoglobin levels below 11 g/dL, whereas SpHb classified only 8.6% in this category. On the upper end of values, CBC classified just 6.9% of observations as having hemoglobin levels between 13-15 g/dL, compared to 38% by SpHb.

To evaluate diagnostic agreement, PPA and NPA were examined across several binary definitions of abnormal hemoglobin (Table 3). PPA values were consistently low: <10% across three of the four defined criteria ((1) Hb <11 g/dL or Hb >15 g/dL: 9.3% (95%CI: 3.3%, 15.2%); (2) Hb <11 g/dL: 7.6% (95%CI: 1.7% - 13.6%); (3) Hb >15 g/dL: 7.7% (95% CI: 0%, 17.4%). PPA was 45.2% (95%CI: 38.5%, 51.8%) for only hemoglobin <11 g/dL or Hb >13 g/dL criteria, which is still <50% in classifying the abnormal anemia state. In contrast, NPA was consistently high, reaching 98.5% for the >15 g/dL threshold and 98.3% for the hemoglobin <11 g/dL alone. This high NPA reflects that the majority of participants had hemoglobin values within the normal range; thus, most negative classifications by SpHb were indeed true negatives. Both Cohen’s kappa and McNemar’s test results (Table S2) showed consistently poor and statistically significant disagreement between SpHb and CBC classifications across all hemoglobin thresholds, further confirming the limited diagnostic concordance between the two methods.

**Table 3.**
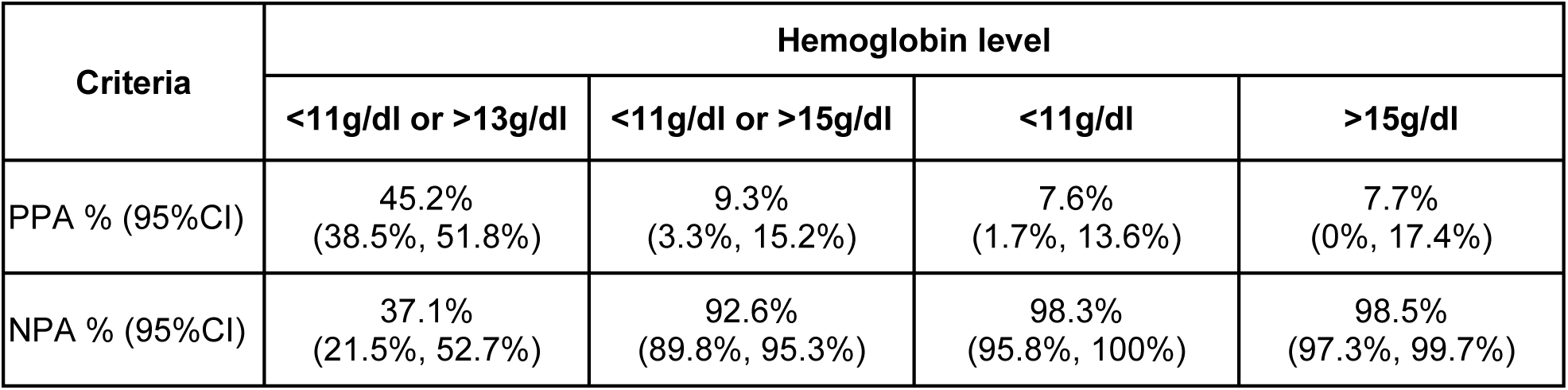
Positive percent agreement (PPA) and negative percent agreement (NPA) between SpHb and CBC on a binary scale.

| Criteria | Hemoglobin level |  |  |  |
| --- | --- | --- | --- | --- |
|  | <11g/dl or >13g/dl | <11g/dl or >15g/dl | <11g/dl | >15g/dl |
| PPA % (95%CI) | 45.2%<br>(38.5%, 51.8%) | 9.3%<br>(3.3%, 15.2%) | 7.6%<br>(1.7%, 13.6%) | 7.7%<br>(0%, 17.4%) |
| NPA % (95%CI) | 37.1%<br>(21.5%, 52.7%) | 92.6%<br>(89.8%, 95.3%) | 98.3%<br>(95.8%, 100%) | 98.5%<br>(97.3%, 99.7%) |

Ordinal agreement across anemia severity categories was assessed using weighted Cohen’s kappa and also showed poor consistency. Kappa values were near zero or negative, indicating no agreement beyond chance across anemia severity categories. Similarly, Harrell’s C-index showed only a 36% concordance (C-index: 0.36±0.01), suggesting that SpHb performed poorly in correctly ranking anemia severity compared to the CBC method.

Evaluating the mean and standard deviation of CBC levels across the different levels of clinical and demographic factors, the CBC levels were notable at 11.8±0.6 among those who were categorized as non-anemic (normal), at 6.0±0.7 among those classified as severely anemic, and at 16.2±1.5 among those who had very high Hb (≥15 g/dL) (Table 4). Additionally, mean CBC levels were highest among women in Zambia at 11.4±1.5 across all measurements, women with higher BMI (overweight and obese) (12.0±1.4 and 12.1±1.4, respectively), among women with a 6-week postpartum measurement (12.1±1.4) and those who were malaria positive (11.0±1.5).

**Table 4.** Mean difference in SpHb and CBC measurements stratified by demographic, health, and nutrition factors.

| Factor | Mean CBC (SD) (g/dL) | Mean Difference<br>(95%CI)<br>(SpHb minus CBC) | Adjusted Mean<br>Difference<sup>2</sup> (95%CI)<br>(SpHb minus CBC) |
| --- | --- | --- | --- |
| Maternal age at enrollment |  |  |  |
| ≥18 years | 11.0 (1.4) | Ref. | Ref. |
| <18 years | 11.0 (1.6) | 0.27 (-0.09, 0.63) | 0.13 (-0.16, 0.41) |
| Tobacco, Betelnut or Smoke |  |  |  |
| No | 10.6 (1.5) | Ref. | Ref. |
| Yes | 11.0 (1.7) | 0.04 (-0.09, 0.18) | -0.18 (-0.29, -0.07) |
| BMI |  |  |  |
| Underweight (<18.5 kg/m<sup>2</sup>) | 11.5 (1.7) | 0.12 (-0.02, 0.26) | -0.06 (-0.06, 0.17) |
| Normal (18.5 - <25 kg/m<sup>2</sup>) | 11.1 (1.7) | Ref. | Ref. |
| Overweight (≥25 - <30 kg/m<sup>2</sup>) | 12.0 (1.4) | -0.15 (-0.26, -0.05) | -0.01 (-0.08, 0.10) |
| Obese (≥30 kg/m<sup>2</sup>) | 12.1 (1.5) | -0.31 (-0.43, -0.18) | -0.07 (-0.18, 0.05) |
| Gestational age at enrollment | Not applicable | 0.02 (0.01, 0.03) | 0.01 (0.00, 0.02) |
| Gestational age by trimester |  |  |  |
| Trimester 1 (≤13 wks) | 11.3 (1.7) | Ref. | Ref. |
| Trimester 2 (>13 to ≤26 wks) | 10.5 (1.3) | 0.92 (0.83, 1.03) | 0.33 (0.24, 0.42) |
| Trimester 3 (>26 to 40 wks) | 10.6 (1.3) | 1.02 (0.94, 1.11) | 0.46 (0.38, 0.54) |
| 6-weeks postpartum | 12.1 (1.4) | -0.13 (-0.25, -0.02) | 0.23 (0.13, 0.33) |
| HIV status at enrollment |  |  |  |
| No | 11.0 (1.5) | Ref. | Ref. |
| Yes | 10.8 (1.9) | 0.08 (-0.05, 0.20) | -0.13 (-0.23, -0.04) |
| Malaria status at any visit |  |  |  |
| No | 11.0 (1.6) | Ref. | Ref. |
| Yes | 10.2 (1.5) | 0.44 (0.24, 0.64) | 0.19 (0.04, 0.34) |
| Site<sup>1</sup> |  |  |  |
| Pakistan | 10.9 (1.6) | Ref. | Ref. |
| Kenya | 10.4 (1.6) | 1.23 (1.14, 1.33) | 1.05 (0.97, 1.13) |
| Zambia | 11.4 (1.5) | 0.33 (0.25, 0.42) | 0.66 (0.59, 0.73) |
| Trimester-specific anemia |  |  |  |
| Severe | 6.2 (0.8) | 2.47 (2.16, 2.77) | 2.44 (2.14, 2.74) |
| Moderate | 9.1 (0.8) | 1.58 (1.49, 1.67) | 1.53 (1.44, 1.62) |
| Mild | 10.4 (0.6) | 0.74 (0.66, 0.82) | 0.71 (0.63, 0.78) |
| Normal | 11.7 (0.6) | Ref. | Ref. |
| High Hb 13-<15 g/dl | 13.6 (0.5) | -1.52 (-1.65, -1.40) | -1.20 (-1.33, -1.08) |
| Very high Hb ≥15 g/dl | 16.3 (1.5) | -4.20 (-4.53, -3.86) | -3.95 (-4.28, -3.62) |
| Plasma Volume (PV) (L) |  |  |  |
| ≥ 2.4 (median) | 11.0 (1.5) | Ref. | Ref. |
| < 2.4 (median) | 11.4 (1.6) | 0.79 (0.71, 0.87) | 0.04 (-0.03, 0.12) |
| Plasma Volume (PV) (L) | Not applicable | 1.15 (1.05, 1.24) | 0.05 (-0.04, 0.14) |
| MUAC |  |  |  |
| >23 cm | 11.1 (1.5) | Ref. | Ref. |
| ≤23 cm | 10.7 (1.7) | 0.17 (-0.07, 0.27) | -0.06 (-0.14, 0.01) |
| Ferritin at week 20 and/or 6-week postpartum |  |  |  |
| Normal ($\geq 70$ ug/L)<br>Deficient ($< 70$ ug/L) | 11.5 (1.41)<br>0.5 (1.50) | Ref.<br>0.61 (0.50, 0.71) | Ref.<br>-0.02 (-0.10, 0.06) |

<sup>1</sup>Modeled as fixed effects; CBC measurements for Kenya and Zambia adjusted for altitude
<sup>2</sup>Models are adjusted for gestational age, HIV status, hemoglobin level, plasma volume, and MUAC, except the model for BMI is not adjusted for MUAC and the model for anemia status is not adjusted for hemoglobin level.

Lastly, linear mixed model results assessed predictors of the difference between SpHb and CBC measurements (Table 4). Reported use of tobacco, betelnut, or smoking (β-0.18; 95% CI-0.29,-0.07) and HIV status (β-0.13; 95% CI-0.23,-0.04) were significantly associated with lower SpHb values compared to CBC, even after adjusting for confounders. Compared to the first trimester, gestational age in the second (β=0.33; 95%CI 0.24, 0.42) and third trimesters (β=0.46; 95% CI 0.38, 0.54) and at six weeks postpartum (β=0.23; 95% CI 0.13, 0.33) was also linked to larger SpHb-CBC mean differences when compared to the first trimester even after adjusting for confounders. This pattern was also observed in Kenya (β=1.05; 95% CI: 0.97, 1.13) and Zambia (β=0.66; 95%CI 0.59, 0.73) when compared to Pakistan. Trimester-specific maternal anemia severity as defined by CBC hemoglobin was significantly associated with a mean difference in SpHb and CBC among all levels. Among women with mild (β=0.77; 95%CI 0.63, 0.78), moderate (β=1.53; 95%CI 1.44, 1.62), and severe (β=2.44; 95%CI 2.14, 2.74) anemia, SpHb levels were significantly higher compared to women without anemia after adjusting for MUAC and GA. These differences were also statistically significant but with higher CBC levels than SpHb hemoglobin levels among women with high Hb of 13-<15 g/dL (β=-1.20; 95%CI-1.33,-1.08) and very high Hb ≥15 g/dl (β=-3.95; 95%CI-4.28,-3.62).

## DISCUSSION

The study presents a comprehensive assessment of the Masimo Total Hemoglobin SpHb device’s efficacy in measuring hemoglobin levels among pregnant and postpartum women in Zambia, Kenya, and Pakistan. Our results indicate that the Masimo SpHb device consistently yields higher hemoglobin readings compared to the gold standard CBC method, with greater differences at very low (<7.0 g/dL) and very high (≥15 g/dL) hemoglobin values. As such, the Masimo SpHb device tended to overestimate normal and high hemoglobin levels while underestimating low hemoglobin levels indicative of anemia as defined by CBC. The various analytical approaches highlighted significant statistical and clinical differences between SpHb and CBC methods. These findings underscore the importance of further validation and refinement of non-invasive hemoglobin measurement devices, especially in pregnant populations where accurate diagnosis of anemia is critical for timely intervention to improve maternal and fetal health outcomes.

Our results align with recent studies on the diagnostic accuracy of the point-of-care SpHb device as compared to laboratory CBC for identifying anemia in pregnant women. A large study of pregnant women in Kenya found that the Masimo Rad-67 Pulse CO-Oximeter overestimates low hemoglobin values measured by CBC and underestimates higher hemoglobin values, resulting in low sensitivity for detecting pregnant women with anemia (18.6%) [19]. In Canada, a study among postpartum women also found that SpHb measured by the Rad-67 overestimated hemoglobin on average compared to the gold standard, with a bias of +2.2 (±1.1) g/dL on the Bland-Altman plot, and a sensitivity of 16% for detecting postpartum anemia [20]. Finally, a small study among women on the day of delivery found a similarly low sensitivity (20%) for the diagnosis of anemia [27]. This body of evidence suggests that the SpHb device requires improvements before it can confidently be used for clinical anemia diagnoses during pregnancy.

Our findings suggest that the limited accuracy of the Masimo SpHb device among a pregnant population may be partially explained by the physiological and sociodemographic factors associated with pregnancy. Our risk factor analyses revealed that SpHb readings were significantly less accurate among women who used tobacco, betelnut or smoked and among women living with HIV [28, 29]. These exposures may influence peripheral perfusion due to the endothelial cell dysfunction, which may in turn affect optical signal quality used by pulse CO-oximetry [30]. In addition, the discrepancy between SpHb and CBC values increased with advancing gestational age, suggesting that pregnancy-related physiological changes, such as plasma volume expansion may impair device accuracy [31]. Differences by country were also observed, with greater discrepancies in Kenya and Zambia compared to Pakistan, possibly reflecting regional environmental, nutritional or health system differences. Our results further demonstrate systematic bias of the Masimo device across the hemoglobin spectrum, which is particularly problematic in antenatal care settings where accurately detecting both low and high extremes of hemoglobin is clinically critical.

Strengths of this study include its large sample size and data collection in two distinct regions, (sub-Saharan Africa and South Asia), three countries (Pakistan, Kenya, and Zambia), and over five time points in pregnancy and postpartum. The robust overall data completeness enabled statistical power for both continuous and categorical analyses. Across multiple analytical methods, significant and clinically relevant discrepancies were consistently observed between hemoglobin measurements and anemia detection by the Masimo SpHb device compared to the gold standard CBC testing. This study also presented a few limitations. The comparison was limited to the Masimo Rad-67 SpHb device and therefore may not be generalizable to other devices which may measure non-invasive Hb. Furthermore, our analysis focused on measurement accuracy and anemia classification but did not evaluate how use of the Masimo SpHb device might influence clinical decision-making in real-world settings.

The comparison of noninvasive hemoglobin measurements using the Masimo SpHb with the gold standard CBC hemoglobin values across Kenya, Zambia, and Pakistan offers valuable insight into the feasibility and performance of point-of-care technologies in diverse antenatal care settings. Given the high likelihood of missing a diagnosis of maternal anemia, our results do not support the use of SpHb devices to measure hemoglobin in pregnant or postpartum women. Future research is needed to refine SpHb measurement algorithms for pregnant populations, with a focus on enhancing accuracy at both low and high hemoglobin levels, where precise identification is most crucial. Until greater accuracy is demonstrated in this population, confirmatory testing with laboratory-based methods remain essential for guiding clinical decision-making during this period

## ETHICAL APPROVALS

Approvals were received for the PRISMA Maternal and Newborn Health Study from the following IRBs and ERCs in each country: Kenya (KEMRI Scientific and Ethics Review Unit: KEMRI/SERU/4166; Liverpool School of Tropical Medicine Ethics Committee: 23-020 Jaramogi Oginga Odinga Teaching and Referral Hospital (JOOTRH) Institutional Scientific Ethics Review Committee ISERC: ISERC/JOOTRH/549/2024); Pakistan (The Aga Khan University ERC: No. 2023-9100-26333; Pakistan National Institutes of Health—Health Research Institute, National Bioethics Committee: 4-87/NBCR-1023/23/973); Zambia (University of North Carolina Chapel Hill Office of Human Research Ethics: Study No. 14-2113; University of Zambia Biomedical Research Ethics Committee: 016-04-14); and the United States (The George Washington University IRB: NCR224396).

## FINANCIAL DISCLOSURE

This work is supported by the Gates Foundation (grant numbers: INV-003601 to Victor Akelo; INV-005776 to Zahra Hoodbhoy; INV-057218 to Margaret Kasaro; INV-041999 to Emily R. Smith).

M. Bridget Spelke is supported by K01TW012426 from NIH/FIC. The funders had no role in study design, data collection and analysis, decision to publish, or preparation of the manuscript.

## Data Availability

Data collection for the parent study (i.e. PRISMA MNH) is ongoing. Upon the conclusion of the study, the consortium may determine to make datasets publicly available. Requests for limited interim datasets will be considered.

## ACKNOWLEDGEMENTS

The authors would like to thank the members and study participants of the Pregnancy Risk, Infant Surveillance, and Measurement Alliance (PRISMA) for their time and efforts. This work was supported by the Gates Foundation. The conclusions and opinions expressed in this work are those of the author(s) alone and shall not be attributed to the Foundation. Under the grant conditions of the Foundation, a Creative Commons Attribution 4.0 License has already been assigned to the Author Accepted Manuscript version that might arise from this submission. Please note works submitted as a preprint have not undergone a peer review process.

## COLLABORATOR STATEMENT

The Pregnancy Risk, Infant Surveillance, and Measurement Alliance (PRISMA) Consortium includes the following institutions and collaborators: Aga Khan University [Amna Khan, Asad Sheikh, Fyezah Jehan], Vital Pakistan Trust [Kinza Farooqui, Farzana Shaheen, Karim Jivani], Kenya Medical Research Institute [Nancy Musimbi, Joab Kuya, Nelson Mandela, Linda Ogolla, Felix Hayara, Irene Migot, Charles Odondi, Lydia Ojowi, John Miguya, Joyce Were, Zaccheus Were, Kephas Otieno, Cynthia Ogwang, Maryanne Nyajom, Thomas Misore], University of North Carolina at Chapel Hill [Felistas Mbewe, Joni Price, Jeffrey Stringer, Bethany Freeman], The George Washington University [Erin Oakley, Jaime Marquis, Jamie Minchin, Jennifer Seager, James Tielsch, Precious Williams, Sarah Tritsch, Savannah O’Malley, Stacie Loisate, Xinyi Li, Nazia Binte Ali], and the Harvard T.H. Chan School of Public Health [Christopher R. Sudfeld].

## SUPPLEMENTARY MATERIAL

**Table S1.** Total number of SpHb and CBC measurements at each visit at each site among women with a visit.

| Site | Timepoint | SpHb (%) | CBC (%) | SpHb and CBC (%) |
| --- | --- | --- | --- | --- |
| Pakistan | ANC<20 | 897 (99.7) | 900 (100.0) | 897 (99.7) |
|  | ANC20 | 768 (95.4) | 769 ( 95.5) | 756 (93.9) |
|  | ANC28 | 762 (87.7) | 747 ( 86.0) | 723 (83.2) |
|  | ANC36 | 616 (82.4) | 627 ( 83.8) | 568 (75.9) |
|  | PNC6 | 753 (88.0) | 547 ( 63.9) | 529 (61.8) |
| Kenya | ANC<20 | 881 (97.9) | 899 (99.9) | 881 (97.9) |
|  | ANC20 | 605 (81.1) | 464 (62.2) | 365 (48.9) |
|  | ANC28 | 608 (68.1) | 528 (59.1) | 319 (35.7) |
|  | ANC36 | 459 (52.1) | 491 (55.7) | 193 (21.9) |
|  | PNC6 | 291 (34.7) | 493 (58.8) | 74 ( 8.8) |
| Zambia | ANC<20 | 873 (97.0) | 896 (99.6) | 869 (96.6) |
|  | ANC20 | 657 (88.8) | 658 (88.9) | 654 (88.4) |
|  | ANC28 | 788 (97.8) | 788 (97.8) | 782 (97.0) |
|  | ANC36 | 684 (99.3) | 674 (97.8) | 669 (97.1) |
|  | PNC6 | 672 (83.8) | 677 (84.4) | 661 (82.4) |

ANC: Antenatal care (<20 weeks, 20 weeks, 28 weeks, 36 weeks gestation)
PNC: Postnatal care (6 weeks postpartum)

**Table S2.** Agreement using Cohen’s Kappa and McNemar’s test between SpHb and CBC on a binary scale for abnormal hemoglobin levels.

| Criteria | Cohen's Kappa (p-value) | McNemar's Test ($\chi^2$) (p-value) |
| --- | --- | --- |
| Hb<11g/dL or Hb>13g/dL | -0.20 (p=0) | 433.18 (<0.001) |
| Hb<11g/dL or Hb>15g/dL | 0.01 (p<0.001) | 4460.00 (p<0.001) |
| Hb<10g/dL or Hb>13g/dL | -0.12 (p<0.001) | 519.34 (p<0.001) |
| Hb<10g/dL or Hb>15g/dL | 0.00 (p=0.001) | 1457 (p<0.001) |
| Hb<10g/dl | 0.00 (p=0.06) | 1245 (p<0.001) |
| Hb <11g/dl | 0.01 (p<0.001) | 3445 (p<0.001) |
| Hb>13g/dl | 0.08 (p=0) | 1796 (p<0.001) |
| Hb >15g/dl | Not applicable | Not applicable |

---

**Figure S1.**
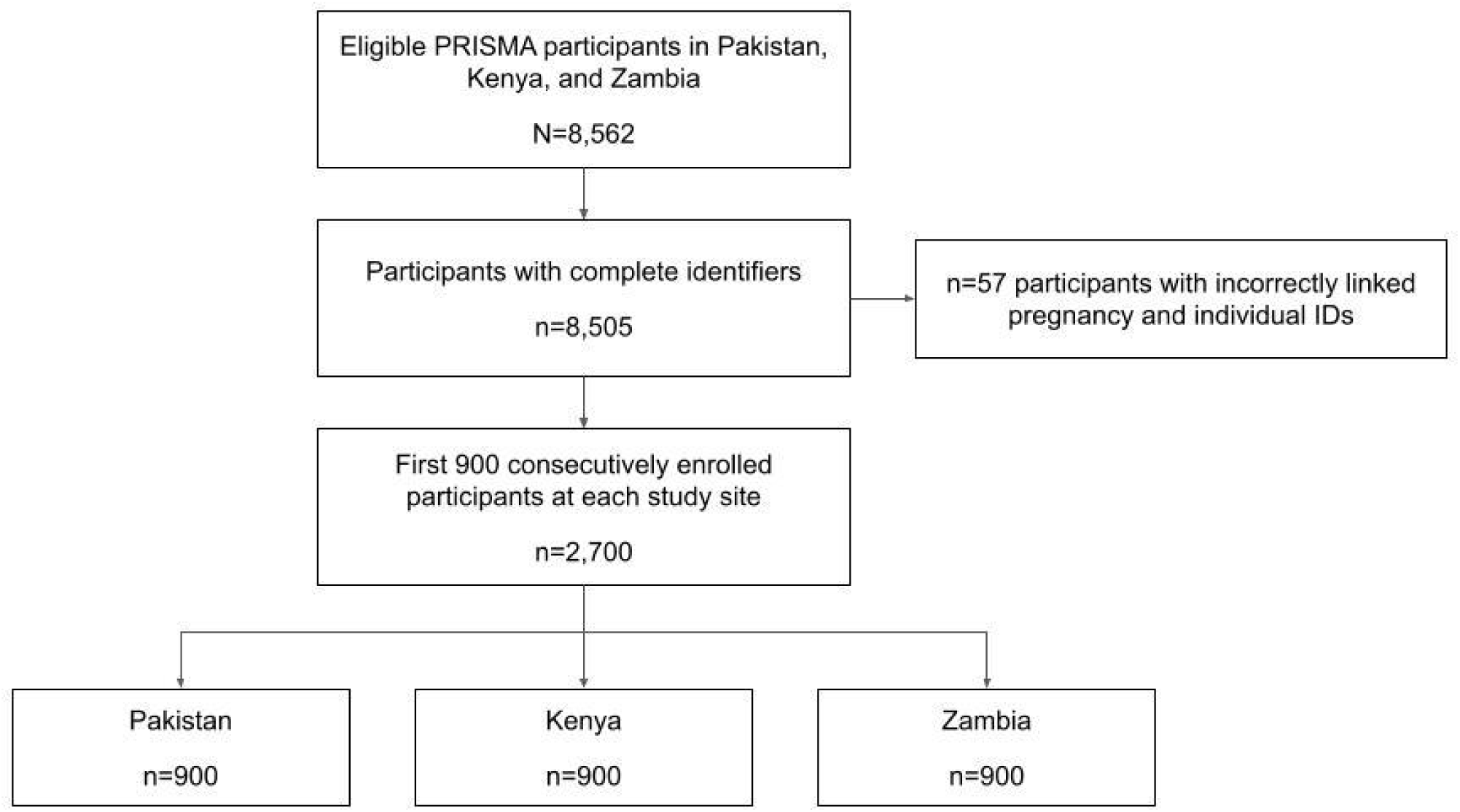
Participant eligibility screening and final sample selection

## Notes

### Competing Interest Statement

The authors have declared no competing interest.

### Clinical Protocols

https://pubmed.ncbi.nlm.nih.gov/37868333/

### Funding Statement

Yes

### Author Declarations

Approvals were received for the PRISMA Maternal and Newborn Health Study from the following IRBs and ERCs in each country: Kenya (KEMRI Scientific and Ethics Review Unit: KEMRI/SERU/4166 Liverpool School of Tropical Medicine Ethics Committee: 23-020 Jaramogi Oginga Odinga Teaching and Referral Hospital (JOOTRH) Institutional Scientific Ethics Review Committee ISERC: ISERC/JOOTRH/549/2024) Pakistan (The Aga Khan University ERC: No. 2023-9100-26333 Pakistan National Institutes of Health—Health Research Institute, National Bioethics Committee: 4-87/NBCR-1023/23/973) Zambia (University of North Carolina Chapel Hill Office of Human Research Ethics: Study No. 14-2113 University of Zambia Biomedical Research Ethics Committee: 016-04-14) and the United States (The George Washington University IRB: NCR224396).

